# A Hybrid AutoML Ensemble Integrating Conventional Learners and Gradient-Boosting Models for Multi-Outcome Prediction in ICU Patients with *Pseudomonas aeruginosa*

**DOI:** 10.1101/2025.06.19.25329970

**Authors:** LV Xiao-chun, Ren Qi, Zhu Lihong, CHEN Kun, WANG Jian-bing, CHEN Fang, JIN Kai-ling, LIN Kai

**Affiliations:** Intensive Care Unit, Zhejiang Hospital, Hangzhou, Zhejiang, China; School of Public Health, Zhejiang University, Hangzhou, Zhejiang, China; Department of Healthcare-associated Infection Management, Zhejiang Hospital, Hangzhou, Zhejiang, China

**Keywords:** Antimicrobial resistance, Pseudomonas aeruginosa, Intensive care unit, Gradient boosting, Automated machine learning

## Abstract

**Background:** Carbapenem resistance in *Pseudomonas aeruginosa* is increasing in intensive care units (ICUs). To enhance antimicrobial stewardship and infection control, we aimed to develop and validate a real-time interpretable hybrid Automated Machine Learning (AutoML) ensemble for multi-outcome prediction.

**Methods:** We retrospectively analyzed 847 adult ICU admissions with *P. aeruginosa* isol ates at a tertiary hospital in Hangzhou, China (January 2018 to December 2024). After a three-stage VTF-MI-L1 feature selection pipeline, XGBoost, LightGBM, CatBoost, random forests, and linear/logistic regression were used as base learners and combined via Bagging, Voting, Stacking, and Gradient Boosting. Nested five-fold cross-validation was used to assess model performance (AUC for classification; MSE, RMSE, MAE, and R^2^ for regression). Interpretability was provided by SHAP values, and the inference latency was recorded.

**Results:** For carbapenem resistance rate (CRR) prediction, the CatBoost regressor (cRMSD = 0.1663; r = 0.8849; R^2^ ≈ 0.78) and the Voting Regressor (cRMSD = 0.1675; r = 0.8838) outperformed all other models (*p* < 0.05). XGB-R achieved the best accuracy and computational efficiency for the last two tests of the CRR of *P. aeruginosa* (CRR-PA-Last2) (*p* < 0.05). In predicting ICU length of stay, XGB-R led with r = 0.9724, cRMSD = 55.7 d, and σ-ratio = 0.88, significantly surpassing Bagging and CatBoost regressors (*p* < 0.05). XGB-R also yielded the lowest composite error for the ICU-to-death interval (cRMSD = 205.1 d; r = 0.7741; σ-ratio = 0.71), again outperforming Bagging and CatBoost *(p* < 0.05). Across all four regression outcomes, XGB-R obtained the lowest average rank (1.95), whereas CatBoost and Voting regressors showed particular strengths in predicting resistance. SHAP analysis identified age, carbapenem exposure intensity, and duration of mechanical ventilation and catheterization as the key positive contributors. All top-ranked models required < 50 ms per inference, meeting the bedside real-time requirements.

**Conclusions:** The proposed hybrid AutoML ensemble delivered highly accurate, interpretable, and millisecond-level predictions of diverse resistance-related outcomes, underscoring its potential for ICU antimicrobial stewardship and infection control. Multicenter prospective studies are warranted to confirm the generalizability of these findings.

## 1 Introduction

Antimicrobial resistance (AMR) is a phenomenon in which target pathogenic microorganisms (e.g., bacteria, fungi, viruses, or parasites) develop resistance to antimicrobial agents because of genetic mutations, selection pressures, or changes in biological adaptation mechanisms, leading to the failure of standard treatment regimens [1, 2]. AMR greatly complicates the clinical management of infectious diseases, prolongs hospital stays, increases healthcare costs, and increases mortality rates [3]. The rapid emergence and global dissemination of multidrug-resistant organisms (MDROs)—often termed “superbugs”—have posed serious challenges to healthcare systems and public health globally [4, 5]. Excessive and irrational use of antimicrobial agents is the primary driver of AMR development and dissemination [6].

Intensive care units (ICUs), which are crucial hospital departments treating critically ill patients, have an antimicrobial usage intensity more than tenfold that of general wards. Over 70% of ICU patients receive antimicrobial therapy during hospitalization, often involving combinations or sequential use of multiple antimicrobial agents [7–9]. Consequently, approximately 35% of ICU patients develop MDRO infections and nearly half of these infections are classified as healthcare-associated infections (HAIs) [10].

*Pseudomonas aeruginosa* (*P. aeruginosa*) is one of the most challenging multidrug-resistant pathogens in HAIs, notably posing significant risks within ICUs [11, 12]. Data from the Gulf Cooperation Council countries indicate that resistance rates of *P. aeruginosa* to carbapenems exceed 30%, and the 30-day mortality rate among infected patients is as high as 68.8%. The World Health Organization (WHO) has also listed carbapenem-resistant *P. aeruginosa* (CRPA) as a “critical priority” pathogen, making its control and treatment among the most urgent issues in the management of global AMR [13].

The rapid advancement of artificial intelligence (AI) has facilitated the development of clinical prediction tools with high accuracy and good generalizability [14]. Nevertheless, existing clinical AI research predominantly focuses on traditional clinical endpoints such as mortality, readmission rates, and length of hospital stay. For instance, Olang et al. reviewed 400 ICU studies and identified that only 16 employed AI methodologies, all of which solely utilized mortality as the outcome measure, with a maximum reported area under the curve (AUC) of 0.929; however, AMR was not considered [15]. Similarly, a large-scale single-center investigation only emphasized length of ICU stay and mortality outcomes without addressing the risk of AMR [16]. Broader quality assessments also reflect this trend: among 338 clinical AI models evaluated by de Hond et al., 67% targeted mortality, hospitalization duration, or readmission rates, while research addressing drug-related risks, particularly AMR, was described as extremely scarce [17]. Conversely, two recent systematic reviews targeting AMR prediction models included only 21 to 27 original studies, typically characterized by limitations such as small sample sizes, single-center studies, offline genomic validation only, absence of external validation, and lack of bedside integration [18, 19]. Even in antimicrobial stewardship scenarios, among 18 AI-supported antimicrobial stewardship program (AI-ASP) tools evaluated by Pinto-de-Sá et al., which reported AUC values ranging from 0.64 to 0.992, functionalities were primarily restricted to prescription auditing or inappropriate medication detection. Therefore, the capacity for AMR risk prediction and meaningful clinical translation remains limited [20].

Given these limitations, this study aimed to introduce an innovative multiscale framework to predict AMR risks. The proposed framework integrates Intermediate Feature Fusion with Automated Machine Learning (AutoML) algorithms, to enable precise AMR risk prediction and highly interpretable outputs for ICU patients, ultimately improving the clinical needs of antimicrobial stewardship.

## 2 Materials and methods

### 2.1 Data

#### 2.1.1 Data description

This retrospective study analyzed the clinical data of patients admitted to our hospital’s ICU from January 1, 2018, to December 31, 2024. Complete medical records of 3,293 patients were collected. The following patients were included: (1) those aged ≥18 years; (2) with an ICU stay of at least 24 h; and (3) at least one specimen (blood, urine, secretions, or excretions) positive for *P. aeruginosa* during admission. We excluded patients with missing clinical data. Ultimately, 847 patients met the inclusion criteria.

Collected variables included the following:

1. Demographic information: Age, sex, and length of ICU stay (ICU-LOS).
2. Comorbidities and medical history: Number of pre-diagnosed comorbidities at admission [21].
3. Surgical and invasive procedures: The total number of surgical procedures performed during hospitalization, duration of central venous catheterization (CVC-days), duration of mechanical ventilation (MV-days), and duration of urinary catheterization (UC-days).
4. Antimicrobial prescriptions: Type, dose, administration route, and treatment duration.
5. Microbiological culture results: Microbiological reports from all specimens collected during hospitalization.
6. Antimicrobial susceptibility testing (AST): AMR definitions followed the guidelines of the WHO and the National Health Commission of China [1, 22]. AST procedures strictly adhered to the Clinical and Laboratory Standards Institute (CLSI) and European Committee on Antimicrobial Susceptibility Testing protocols, utilizing the minimum inhibitory concentration to determine resistance.
7. Outcome measures: Including antimicrobial susceptibility of the last isolate of *P. aeruginosa* before discharge (LastPaAST, classified as resistant [R] or susceptible [S]), in-hospital mortality (IHM), ICU length of stay (ICU-LOS), time from ICU admission to death (ICU-Death interval), average carbapenem resistance rate of the last two isolates (CRR-PA-Last2), and carbapenem resistance rate (CRR).

#### 2.1.2 Indicator calculation and data preprocessing

##### (1) Resistance rate calculation

CRR serves as the primary indicator for assessing AMR risk. The range of carbapenems was defined according to the CLSI standards (CLSI M100) [23]. The CRR of *P. aeruginosa* was calculated using Equation:

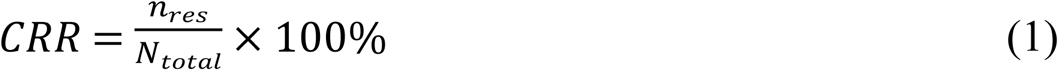

where *n*_*res*_ represents the number of isolates resistant to at least one carbapenem and *N*_*total*_ is the total number of isolates tested.

##### (2) Anatomical Therapeutic Chemical/defined daily dose Calculation

Antimicrobial usage was standardized using the Anatomical Therapeutic Chemical (ATC) classification system and a defined daily dose (DDD) methodology [24]. Standardization was quantified using the ATC/DDD per day of therapy (DOT), calculated as follows:

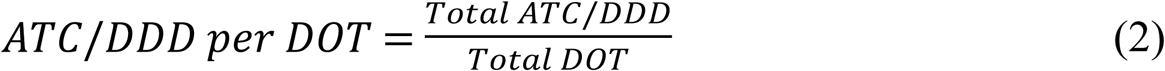

Here, ATC/DDD represents the defined daily dose and DOT refers to the number of days of therapy.

### 2.2 Data preprocessing and feature selection

ICU datasets often exhibit high dimensionality, noise, and multicollinearity, which complicate the stable identification of key features [25]. As shown in Fig 1, we propose a three-stage feature selection method called VTF-MI-L1. Initially, near-zero variance features were removed using a Variance Threshold Filter (VTF). Next, Mutual Information (MI) identified nonlinear relationships, preserving the candidate features with maximal contributions. Finally, L1 regularization generated sparse solutions, addressing multicollinearity and enhancing interpretability [26, 27]. This process reduced the number of feature dimensions from 93 to 48 without compromising the predictive performance.

**Fig 1.**
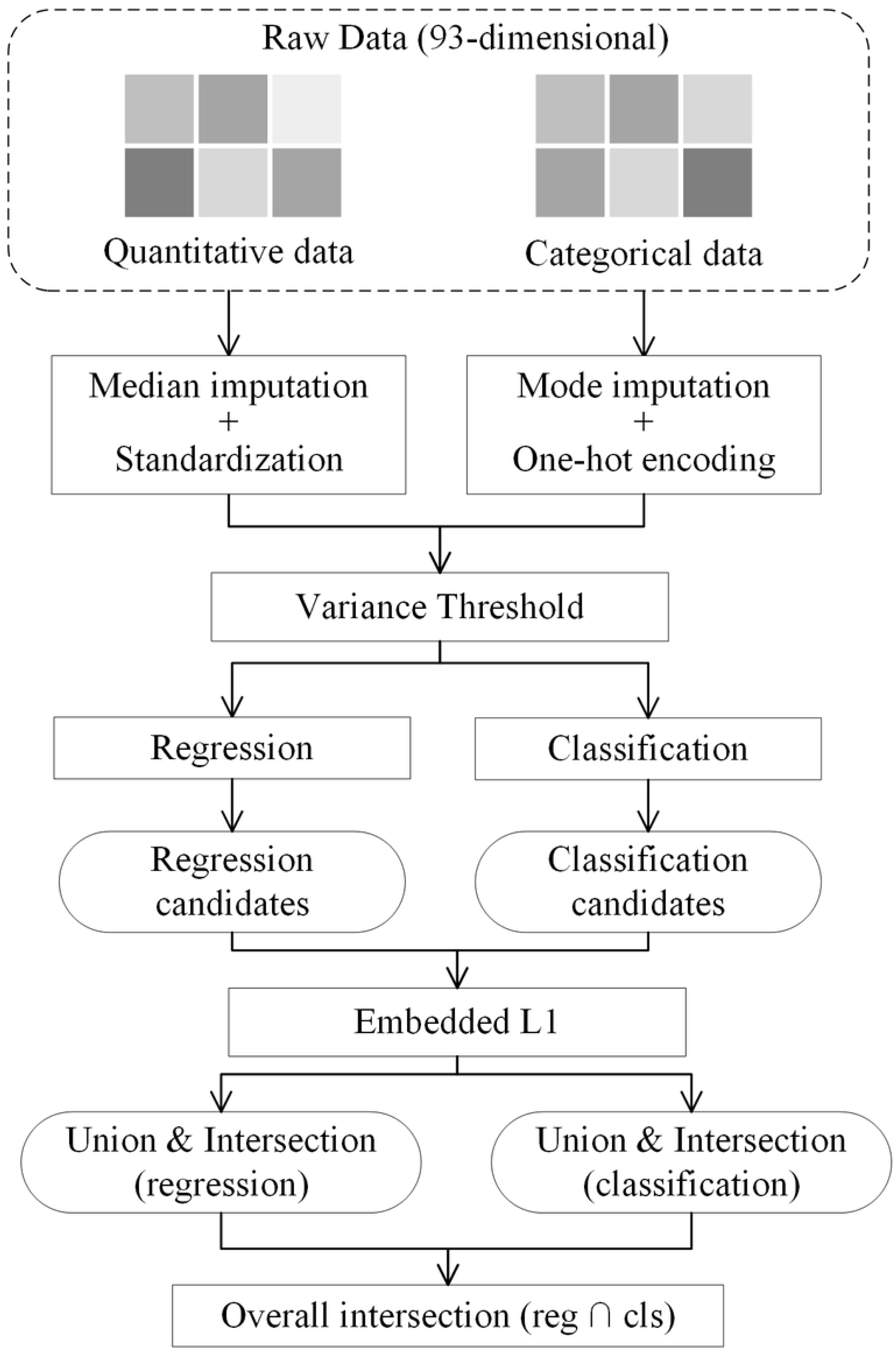
Feature selection flowchart.

First, duplicate records were removed and variables were categorized into numerical and categorical types. Missing numerical data were imputed using median values, whereas mode imputation was used for categorical variables. Numerical variables underwent Z-score normalization [28], and categorical variables were encoded using one-hot encoding [29]. These preprocessing steps were implemented using ColumnTransformer to produce dense feature matrices for modeling. Second, a VTF eliminates variables with near-zero variance, thereby eliminating features that lack substantial predictive information. Third, a mutual-information univariate filter was applied. For the regression tasks, Pearson’s correlation coefficients and MI scores were computed while retaining the top-quartile features. For classification tasks, ANOVA F-values and MI scores were used in a similar manner. Fourth, we employed Embedded L1 Regularization. The regression models utilize Least Absolute Shrinkage and Selection Operator (LASSO) regression with cross-validation, retaining nonzero coefficient features. The classification models used logistic regression with an L1 penalty. Finally, the regression and classification feature sets from these steps were subjected to union and intersection operations, producing robust and comprehensive feature combinations for modeling.

### 2.3 Design and implementation of AutoML

To systematically evaluate the predictive performance, an AutoML framework was developed for regression and classification tasks (Fig 2). The design integrated diverse ensemble strategies and robust cross-validation to enhance prediction accuracy and generalization. For post-standardized preprocessing, the model-building module utilized various base learners as follows: for classification, XGBoost Classifier (XGB-C), LightGBM Classifier (LGB-C), CatBoost Classifier (CAT-C), Random Forest Classifier (RF-C), and logistic regression (LR); for regression, XGBoost Regressor (XGB-R), CatBoost Regressor (CAT-R), LightGBM Regressor (LGB-R), Random Forest Regressor (RF-R), Linear Regression (LR), and Voting Regressor (VR).

**Fig. 2.**
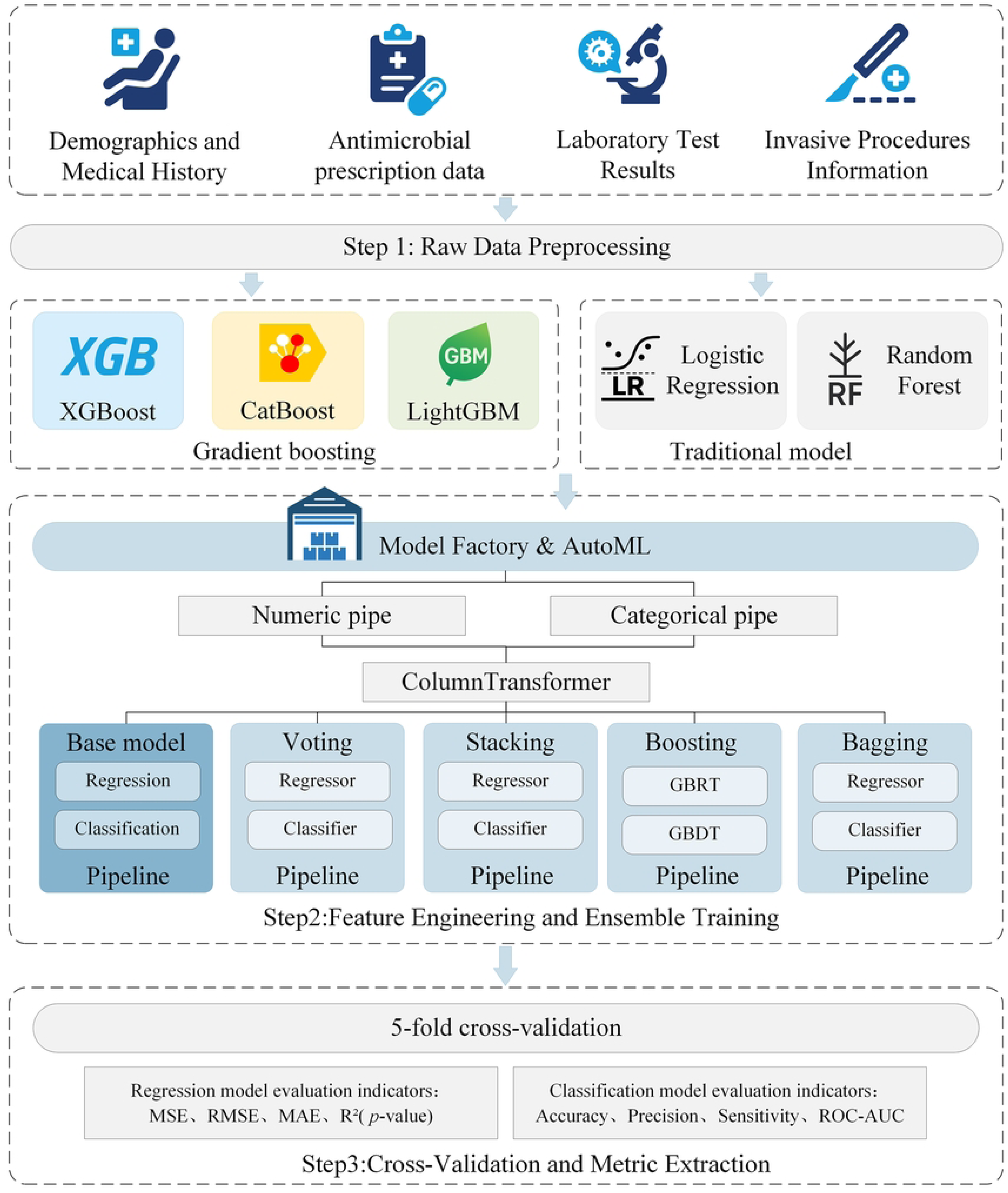
Model architecture framework diagram.

Four mainstream ensemble methods were evaluated to enhance model robustness and generalization:

Bagging: a parallel ensemble method in which multiple base learners are trained on bootstrap samples drawn with replacements from the original training set. The final prediction is calculated by averaging the outputs of the regression tasks or majority voting for the classification tasks. For the bagging regression model (BR), the ensemble prediction *y^^^*_*bag*_ is calculated using (3):

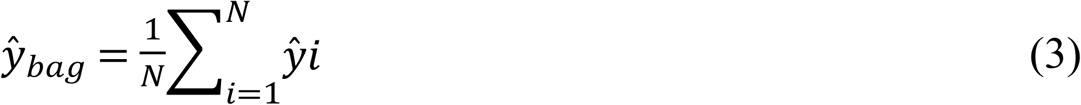

where *N* represents the number of base models, and *y^^^i* denotes the prediction of the *i*-th model.

For the bagging classification model (BC), the ensemble prediction is described by (4):

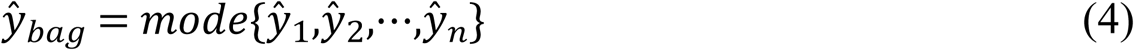

where “*mode*” represents the most frequent predicted class.

Voting: a method in which the predictions of multiple base models are aggregated by weighted averaging for regression or weighted probabilities for classification (soft voting). For the voting regression model (VR), the final prediction was calculated using (5):

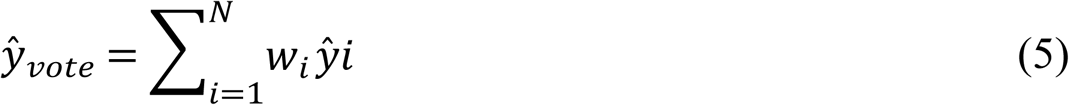

where *w*_*i*_ is the weight assigned to the *i*-th model. For the voting classification model (VC), the class probabilities are aggregated and the predicted class is described by (6):

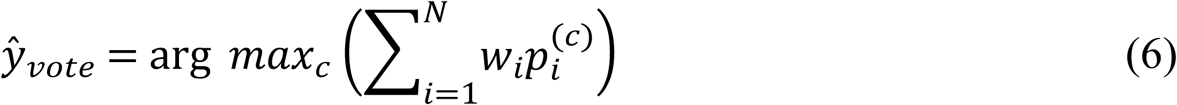

where *p*^(*c*)^_i_ represents the predicted probability of class *c* by the *i*-th model.

Stacking: A hierarchical ensemble method in which predictions from multiple base models are used as new features for a meta-learner. The final prediction is described by (7):

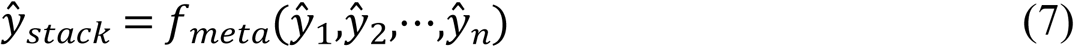

where *f*_*meta*_ represents the metamodel trained on the out-of-fold predictions of the base models.

In this study, stacking was implemented in two forms—the stacking classifier (SC) and the stacking regressor (SR)—corresponding to classification and regression tasks, respectively.

Boosting: A sequential ensemble method in which each new base learner is trained to fit the residuals or gradients of the preceding learners. The ensemble prediction after *m* iterations is represented by Equation (8):

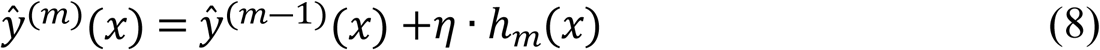

where *y^^^*^(*m*)^(*x*) represents the prediction at iteration *m*, η denotes the learning rate, and *h*_*m*_(*x*) is the newly fitted weak learner.

For regression and classification outcomes, the Boosting component employed two symmetrical implementation strategies:

1. Continuous outcomes (regression): Gradient Boosting Regression Trees (GBRT) were utilized, with the Mean Squared Error (MSE) as the optimization objective. This model effectively captures nonlinear relationships by iteratively fitting residuals, demonstrating superior performance in numerical indicators such as AMR rates.
2. Discrete outcomes (Classification): Gradient Boosting Decision Trees (GBDT) were implemented with log-likelihood as the optimization objective. This approach generates posterior probabilities, facilitating the evaluation of threshold-independent metrics such as the ROC-AUC. These methods were integrated into an end-to-end AutoML pipeline, featuring unified preprocessing and automated base learner mapping to ensemble meta-learners, and managed via the scikit-learn pipeline framework. The models were trained under consistent computing conditions with predefined hyperparameters to minimize variability and ensure reproducibility.

### 2.4 Evaluation methods and statistical analysis

A nested cross-validation framework was employed to systematically evaluate the predictive performance and generalization capability of both the regression and classification models. For regression tasks, 5-fold cross-validation was utilized, with model performance assessed through metrics, including the Mean Absolute Error (MAE), Root Mean Squared Error (RMSE), and Coefficient of Determination (R^2^). The statistical significance of the R^2^ values was assessed using an F-test [30, 31].

Stratified 5-fold cross-validation was adopted for the classification tasks. The evaluated metrics included accuracy, precision, sensitivity, specificity, F1-score, and the Area Under the Receiver Operating Characteristic Curve (ROC-AUC). ROC curves were constructed using out-of-fold predicted probabilities to estimate the AUC.

In the regression tasks, four continuous outcome variables—CRR, CRR-PA-Last2, ICU-Death interval, and ICU-LOS—were analyzed using a unified 5-fold cross-validation procedure, with MSE, RMSE, MAE, and R^2^ calculated for each. Each outcome-metric combination was treated as an independent dataset and eight regression models (XGB-R, LGB-R, CAT-R, RF-R, BR, GBRT, VR, and SR) were ranked. The Friedman test was employed to evaluate the overall differences among these models, with pairwise comparisons subsequently performed using the Nemenyi post-hoc test [32].

In addition, SHapley Additive exPlanations (SHAP) were introduced to interpret models and elucidate the relative importance and directional effects of features on model predictions [33]. To further assess consistency across multiple models and predictive tasks, Taylor diagrams were used to visualize correlations, standard deviations, and Centered Root Mean Square Differences (CRMSD) between predictions and observations.

### 2.5 Statistical methods and modeling tools

All statistical analyses and model development were performed in Python 3.10, utilizing open-source libraries, including Pandas, NumPy, SciPy, scikit-learn, LightGBM, CatBoost, XGBoost, and TensorFlow. Continuous variables are summarized as mean ± standard deviation (SD) if normally distributed, while non-normally distributed data underwent logarithmic transformation [ln(x+1)] and were summarized using geometric means (GM) with geometric standard deviations (GSD). Categorical variables are reported as frequency and percentage (n [%]).

The feature engineering workflow encompasses variance threshold filtering, univariate feature selection (using F-statistics and MI), and embedded feature selection through L1 regularization models (LASSO for regression tasks and logistic regression for classification tasks). Ensemble modeling strategies included Bagging, Voting, Stacking, and Boosting methods, with ridge regression or logistic regression consistently serving as meta-learners for these ensembles. The entire modeling pipeline was automated and standardized using fixed parameter configurations. To statistically compare model performance, regression models were subjected to non-parametric comparisons using the Friedman test, followed by Nemenyi post-hoc tests. Classification models employed bootstrap methods (1,000 repetitions) to compute confidence intervals and two-tailed p-values for pairwise differences in AUC values among models, with false-discovery rate (FDR) correction applied for multiple hypothesis testing. All statistical tests were two-sided, and the significance threshold was set at *p* < 0.05.

## 3 Results

### 3.1 Selected features

A total of 48 feature variables were ultimately included in the analysis based on the feature selection methodology. These features included demographic information, comorbidities, medical history, surgical and invasive procedures, antimicrobial exposure, and microbiological characteristics. A summary of the key feature distributions is presented in Table 1. Among the 847 ICU patients included, 597 were male (70.5%) and 250 were female (29.5%). The cohort was predominantly comprised of older adults, with a median age of 79 years; 77.1% of patients were aged 65 years or older. Regarding surgical history, 427 patients (50.4%) had not undergone surgery, 239 (28.2%) underwent one surgical procedure, and 181 (21.4%) underwent two or more. All non-normally distributed numerical variables were log-transformed prior to statistical analysis, with the results expressed as GM and GSD. The GM durations were 20.28 days (GSD 2.94) for central venous catheterization (CVC-days), 17.86 days (GSD 3.51) for mechanical ventilation (MV-days), and 21.30 days (GSD 3.49) for urinary catheterization (UC-days). The time to the first carbapenem-resistant *P. aeruginosa* (TFC-CRPA) had a GM of 17.76 days (GSD, 2.14). The proportion of initial carbapenem-resistant isolates (IPA-CARBSUSC) was 62.9%. Carbapenems (CARB) showed the highest usage (ATC-DDD GM 6.70, GSD 2.54), followed by antifungals (AF, GM 4.05, GSD 4.10), and β-lactam/β-lactamase inhibitors (BL/BLI, GM 2.35, GSD 2.96). Other agents such as rifampin (RIF), sulfonamides (SUL), penicillin (PEN), and nitrofurantoin (NIT) were used at low levels (GM ≤ 0.01).

**Table 1.**
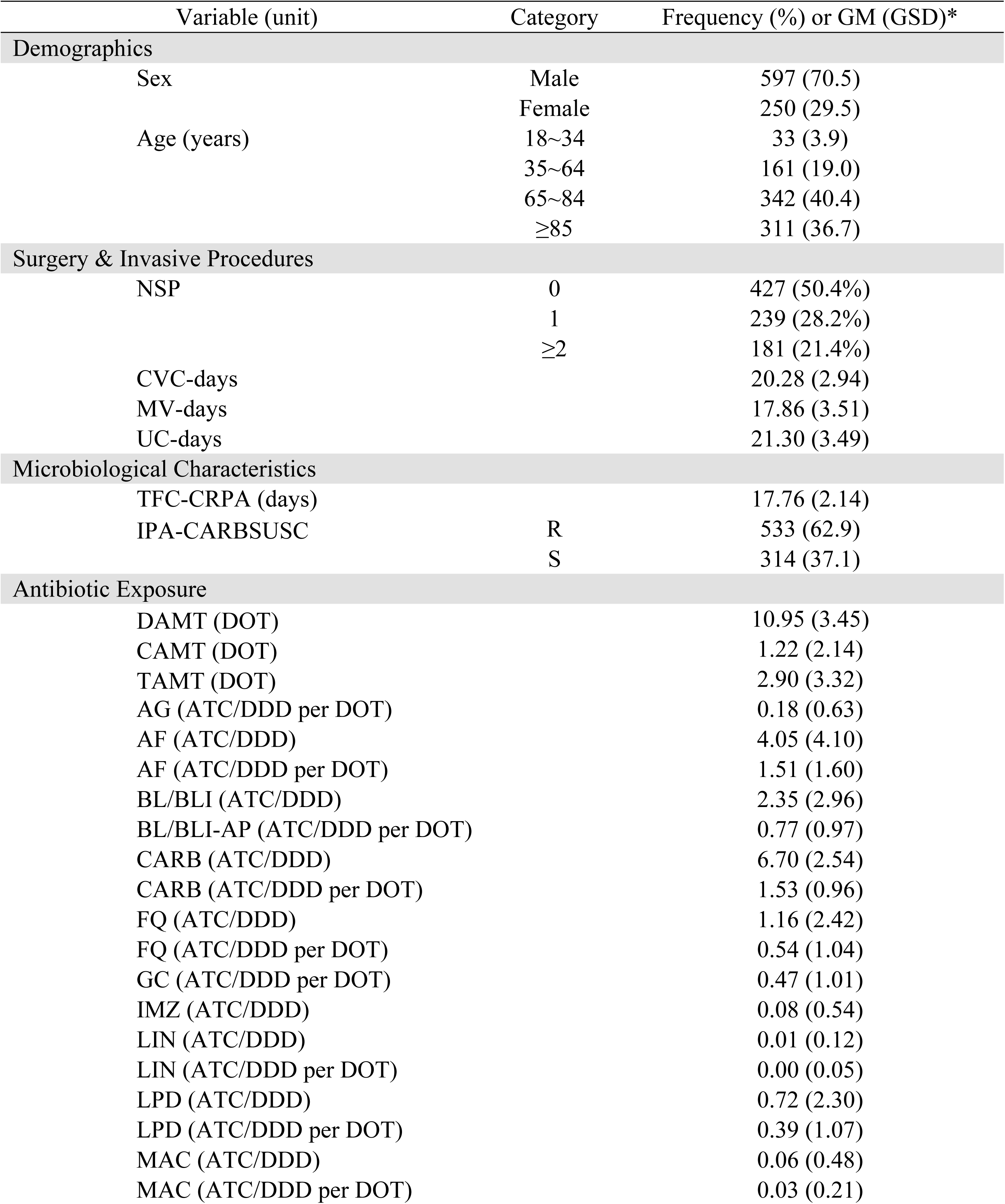
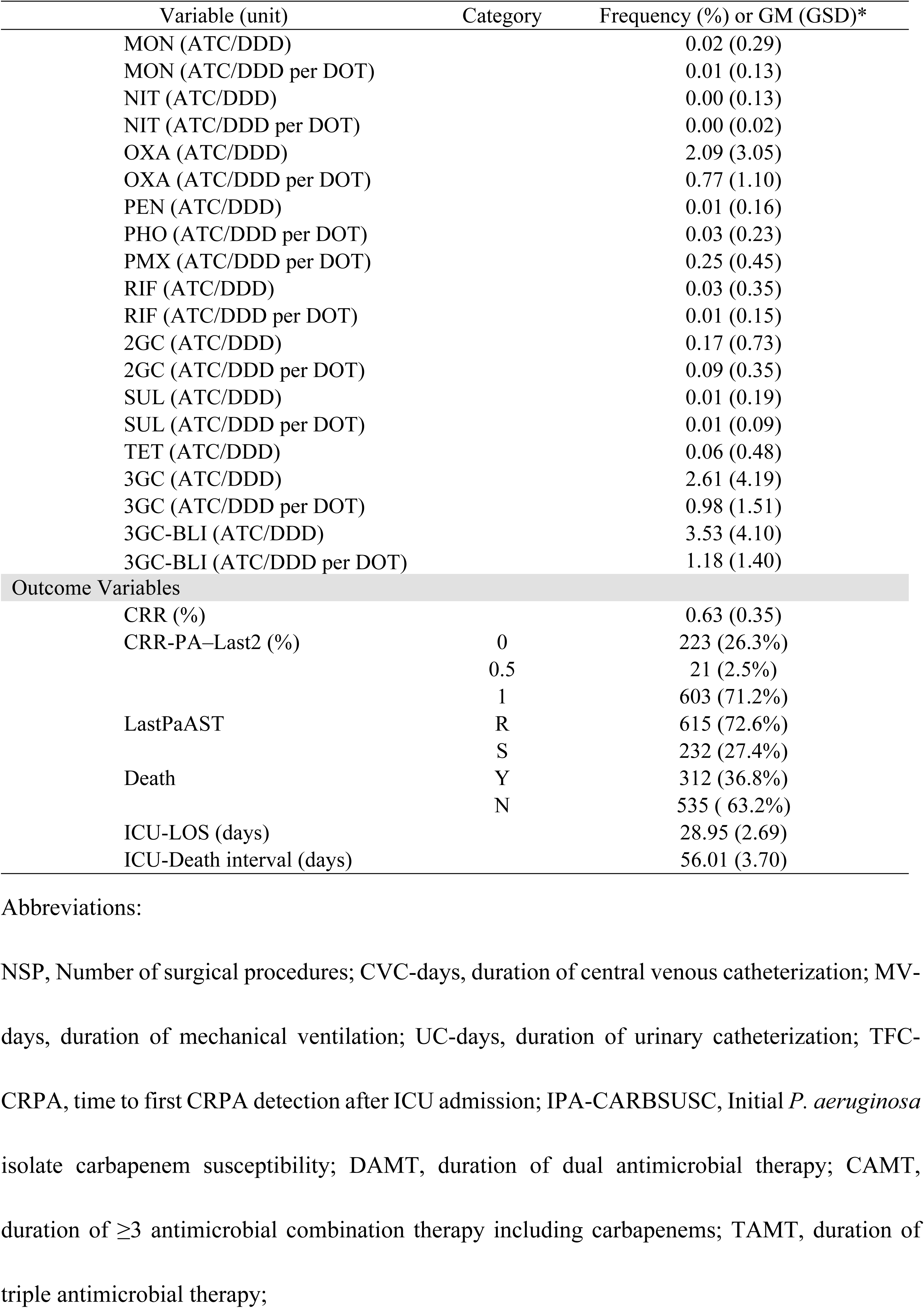
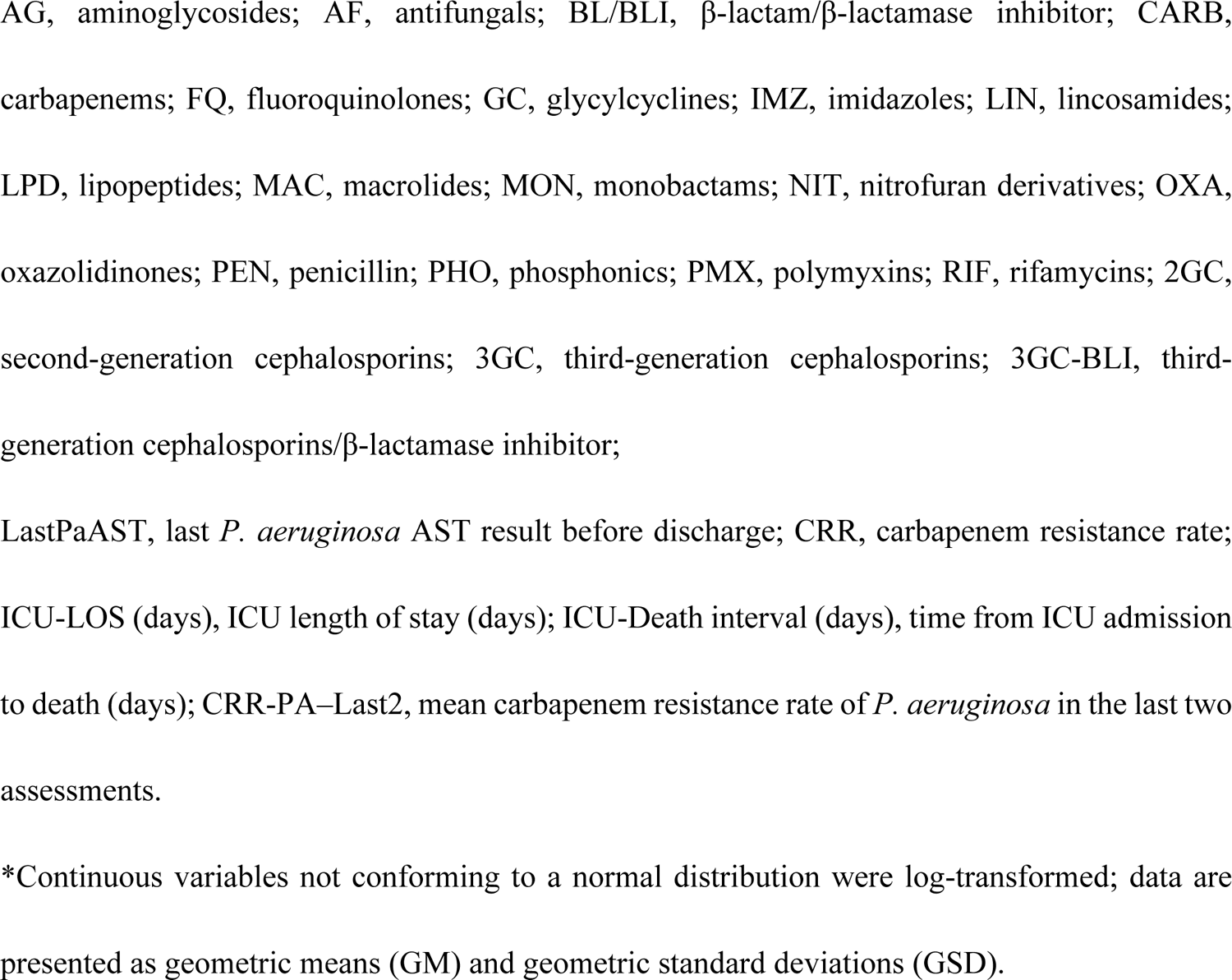
Descriptive statistics for baseline characteristics and outcome variables.

| Variable (unit) | Category | Frequency (%) or GM (GSD)* |
| --- | --- | --- |
| **Demographics** |  |  |
| Sex | Male | 597 (70.5) |
|  | Female | 250 (29.5) |
| Age (years) | 18~34 | 33 (3.9) |
|  | 35~64 | 161 (19.0) |
|  | 65~84 | 342 (40.4) |
|  | ≥85 | 311 (36.7) |
| **Surgery & Invasive Procedures** |  |  |
| NSP | 0 | 427 (50.4%) |
|  | 1 | 239 (28.2%) |
|  | ≥2 | 181 (21.4%) |
| CVC-days |  | 20.28 (2.94) |
| MV-days |  | 17.86 (3.51) |
| UC-days |  | 21.30 (3.49) |
| **Microbiological Characteristics** |  |  |
| TFC-CRPA (days) |  | 17.76 (2.14) |
| IPA-CARBSUSC | R | 533 (62.9) |
|  | S | 314 (37.1) |
| **Antibiotic Exposure** |  |  |
| DAMT (DOT) |  | 10.95 (3.45) |
| CAMT (DOT) |  | 1.22 (2.14) |
| TAMT (DOT) |  | 2.90 (3.32) |
| AG (ATC/DDD per DOT) |  | 0.18 (0.63) |
| AF (ATC/DDD) |  | 4.05 (4.10) |
| AF (ATC/DDD per DOT) |  | 1.51 (1.60) |
| BL/BLI (ATC/DDD) |  | 2.35 (2.96) |
| BL/BLI-AP (ATC/DDD per DOT) |  | 0.77 (0.97) |
| CARB (ATC/DDD) |  | 6.70 (2.54) |
| CARB (ATC/DDD per DOT) |  | 1.53 (0.96) |
| FQ (ATC/DDD) |  | 1.16 (2.42) |
| FQ (ATC/DDD per DOT) |  | 0.54 (1.04) |
| GC (ATC/DDD per DOT) |  | 0.47 (1.01) |
| IMZ (ATC/DDD) |  | 0.08 (0.54) |
| LIN (ATC/DDD) |  | 0.01 (0.12) |
| LIN (ATC/DDD per DOT) |  | 0.00 (0.05) |
| LPD (ATC/DDD) |  | 0.72 (2.30) |
| LPD (ATC/DDD per DOT) |  | 0.39 (1.07) |
| MAC (ATC/DDD) |  | 0.06 (0.48) |
| MAC (ATC/DDD per DOT) |  | 0.03 (0.21) |
| MON (ATC/DDD) |  | 0.02 (0.29) |
| MON (ATC/DDD per DOT) |  | 0.01 (0.13) |
| NIT (ATC/DDD) |  | 0.00 (0.13) |
| NIT (ATC/DDD per DOT) |  | 0.00 (0.02) |
| OXA (ATC/DDD) |  | 2.09 (3.05) |
| OXA (ATC/DDD per DOT) |  | 0.77 (1.10) |
| PEN (ATC/DDD) |  | 0.01 (0.16) |
| PHO (ATC/DDD per DOT) |  | 0.03 (0.23) |
| PMX (ATC/DDD per DOT) |  | 0.25 (0.45) |
| RIF (ATC/DDD) |  | 0.03 (0.35) |
| RIF (ATC/DDD per DOT) |  | 0.01 (0.15) |
| 2GC (ATC/DDD) |  | 0.17 (0.73) |
| 2GC (ATC/DDD per DOT) |  | 0.09 (0.35) |
| SUL (ATC/DDD) |  | 0.01 (0.19) |
| SUL (ATC/DDD per DOT) |  | 0.01 (0.09) |
| TET (ATC/DDD) |  | 0.06 (0.48) |
| 3GC (ATC/DDD) |  | 2.61 (4.19) |
| 3GC (ATC/DDD per DOT) |  | 0.98 (1.51) |
| 3GC-BLI (ATC/DDD) |  | 3.53 (4.10) |
| 3GC-BLI (ATC/DDD per DOT) |  | 1.18 (1.40) |
| Outcome Variables |  |  |
| CRR (%) |  | 0.63 (0.35) |
| CRR-PA–Last2 (%) | 0 | 223 (26.3%) |
|  | 0.5 | 21 (2.5%) |
|  | 1 | 603 (71.2%) |
| LastPaAST | R | 615 (72.6%) |
|  | S | 232 (27.4%) |
| Death | Y | 312 (36.8%) |
|  | N | 535 (63.2%) |
| ICU-LOS (days) |  | 28.95 (2.69) |
| ICU-Death interval (days) |  | 56.01 (3.70) |

Abbreviations:
NSP, Number of surgical procedures; CVC-days, duration of central venous catheterization; MV-days, duration of mechanical ventilation; UC-days, duration of urinary catheterization; TFC-CRPA, time to first CRPA detection after ICU admission; IPA-CARBSUSC, Initial *P. aeruginosa* isolate carbapenem susceptibility; DAMT, duration of dual antimicrobial therapy; CAMT, duration of $\geq 3$ antimicrobial combination therapy including carbapenems; TAMT, duration of triple antimicrobial therapy;
AG, aminoglycosides; AF, antifungals; BL/BLI, β-lactam/ β-lactamase inhibitor; CARB, carbapenems; FQ, fluoroquinolones; GC, glycyclines; IMZ, imidazoles; LIN, lincosamides; LPD, lipopeptides; MAC, macrolides; MON, monobactams; NIT, nitrofurans derivatives; OXA, oxazolidinones; PEN, penicillin; PHO, phosphonics; PMX, polymyxins; RIF, rifamycins; 2GC, second-generation cephalosporins; 3GC, third-generation cephalosporins; 3GC-BLI, third-generation cephalosporins/ β-lactamase inhibitor; LastPaAST, last *P. aeruginosa* AST result before discharge; CRR, carbapenem resistance rate; ICU-LOS (days), ICU length of stay (days); ICU-Death interval (days), time from ICU admission to death (days); CRR-PA-Last2, mean carbapenem resistance rate of *P. aeruginosa* in the last two assessments.
\*Continuous variables not conforming to a normal distribution were log-transformed; data are presented as geometric means (GM) and geometric standard deviations (GSD).

### 3.2 Outcome variables

As shown in Table 1, the primary and secondary outcomes of the study were as follows. The overall *CRR* was 0.63 (SD 0.35). For the CRR-PA-Last2, 223 patients (26.3%) had a value of 0, 21 patients (2.5%) had a value of 0.5, and 603 patients (71.2%) had a value of 1. Regarding the LastPaAST, 615 patients (72.6%) were resistant (R) and 232 patients (27.4%) were susceptible (S). IHM occurred in 312 patients (36.8%), and 535 patients (63.2%) were discharged alive. ICU-LOS was 28.95 days (SD, 2.69), and the ICU-Death interval was 56.01 days (SD, 3.70).

### 3.3 Performance evaluation

#### 3.3.1 Classification tasks

We first generated raw ROC curves for all the classification models based on their probability outputs and used the AUC to evaluate the overall discriminative performance. The sensitivity thresholds were aligned to approximately 0.800 for all the models, after which the threshold-dependent metrics were calculated. We further applied 1,000-fold bootstrapping and Benjamini– Hochberg correction to obtain FDR-adjusted p-values (*q*-values), allowing for a systematic comparison of model performance for two binary outcomes: IHM and LastPaAST.

For the IHM outcome, ROC curves (Fig. 3a) showed that Voting Classifier (VC) achieved the highest AUC (0.842), indicating a superior ranking ability. When evaluating the threshold-dependent performance at a fixed sensitivity, VC outperformed the other models with the highest specificity (0.751), accuracy (0.770), positive predictive value (PPV: 0.653), and negative predictive value (NPV: 0.866), as shown in Table 2. Pairwise AUC comparisons (Table 3) revealed significant improvements of VC over LGB-C (ΔAUC = 0.014, 95% CI: 0.007–0.022, q < 0.05) and XGB-C (ΔAUC = 0.011, 95% CI: 0.005–0.018, q < 0.05), while differences with other models were not statistically significant (q = 0.05).

**Fig. 3.**
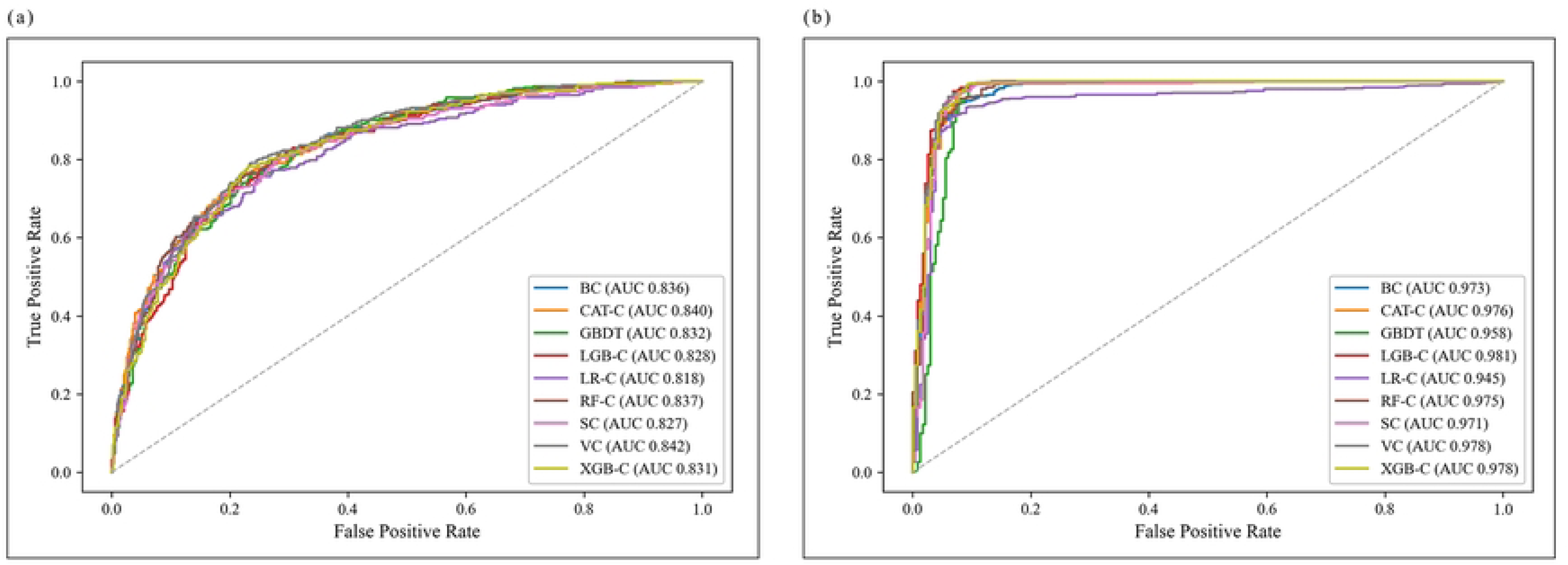
ROC curves of nine classifiers for two binary outcomes. The panels show (a) IHM and (b) LastPaAST. The curves represent the mean five-fold cross-validation, and the AUC values are listed in the legend.

**Table 2.** Discrimination metrics of nine classification models for two binary outcomes.

| Target | Model | AUC | Sens | Spec | Acc | PPV | NPV |
| --- | --- | --- | --- | --- | --- | --- | --- |
| Death | BC | 0.836 | 0.800 | 0.720 | 0.750 | 0.625 | 0.861 |
|  | CAT-C | 0.840 | 0.800 | 0.695 | 0.734 | 0.605 | 0.857 |
|  | GBDT | 0.832 | 0.800 | 0.697 | 0.736 | 0.607 | 0.857 |
|  | LGB-C | 0.828 | 0.800 | 0.716 | 0.747 | 0.622 | 0.861 |
|  | LR-C | 0.818 | 0.800 | 0.652 | 0.707 | 0.573 | 0.849 |
|  | RF-C | 0.837 | 0.800 | 0.723 | 0.752 | 0.628 | 0.862 |
|  | SC | 0.827 | 0.800 | 0.718 | 0.749 | 0.623 | 0.861 |
|  | VC | **0.842** | 0.800 | **0.751** | **0.770** | **0.653** | **0.866** |
|  | XGB-C | 0.831 | 0.800 | 0.733 | 0.758 | 0.636 | 0.863 |
| LastPaAST | BC | 0.973 | 0.800 | 0.961 | 0.844 | 0.982 | 0.645 |
|  | CAT-C | 0.976 | 0.800 | 0.966 | 0.845 | 0.984 | 0.646 |
|  | GBDT | 0.958 | 0.800 | 0.944 | 0.839 | 0.974 | 0.640 |
|  | LGB-C | **0.981** | 0.800 | **0.974** | **0.848** | **0.988** | **0.648** |
|  | LR-C | 0.945 | 0.800 | 0.961 | 0.844 | 0.982 | 0.645 |
|  | RF-C | 0.975 | 0.800 | 0.970 | 0.847 | 0.986 | 0.647 |
|  | SC | 0.971 | 0.800 | 0.961 | 0.844 | 0.982 | 0.645 |
|  | VC | 0.978 | 0.800 | 0.966 | 0.845 | 0.984 | 0.646 |
|  | XGB-C | 0.978 | 0.800 | 0.966 | 0.845 | 0.984 | 0.646 |

The area under the curve (AUC), sensitivity (Sens), specificity (Spec), accuracy (Acc), positive predictive value (PPV), and negative predictive value (NPV) were reported for in-hospital mortality (Death) and last *Pseudomonas aeruginosa* AST (LastPaAST). Statistically significant AUCs are shown in bold.

**Table 3.** Pairwise DeLong comparisons of AUC between classification models with 95% confidence intervals and FDR-adjusted q-values.

| Target | Model 1 | Model 2 | ΔAUC 95% CI,<br>Lower | ΔAUC 95% CI,<br>Upper | *q* |
| --- | --- | --- | --- | --- | --- |
| Death | VC | XGB-C | 0.005 | 0.018 | **0.000** |
|  | LGB-C | VC | -0.022 | -0.007 | **0.000** |
|  | BC | VC | -0.018 | 0.005 | 0.420 |
|  | CAT-C | VC | -0.009 | 0.003 | 0.503 |
|  | GBDT | VC | -0.023 | 0.000 | 0.129 |
|  | RF-C | VC | -0.015 | 0.006 | 0.503 |
|  | LR-C | VC | -0.045 | -0.004 | 0.058 |
|  | SC | VC | -0.032 | 0.000 | 0.128 |
| LastPaAST | BC | LGB-C | -0.012 | -0.004 | **0.000** |
|  | CAT-C | LGB-C | -0.009 | 0.000 | 0.122 |
|  | GBDT | LGB-C | -0.039 | -0.009 | **0.000** |
|  | LGB-C | LR-C | 0.021 | 0.053 | **0.000** |
|  | LGB-C | RF-C | 0.002 | 0.010 | **0.010** |
|  | LGB-C | SC | 0.002 | 0.020 | **0.025** |
|  | LGB-C | XGB-C | 0.001 | 0.006 | **0.030** |
|  | LGB-C | VC | -0.002 | 0.008 | 0.330 |

ΔAUC denotes the difference in AUC (Model 1 – Model 2); CIs are two-sided 95% intervals. The *q* value was the FDR-adjusted *p* value after Benjamini–Hochberg correction, and the difference was considered statistically significant only when $q < 0.05$ .

For the LastPaAST outcome (Fig. 3b), the ROC curve of LGB-C was closest to the top-left corner, yielding an AUC of 0.981. VC and XGB-C closely followed (AUC = 0.978), whereas CAT-C and RF-C had AUCs values of 0.976 and 0.975, respectively (Table 2). At a fixed sensitivity of 0.800, LGB-C achieved the best threshold-dependent metrics, with a specificity of 0.974, an accuracy of 0.848, and a PPV of 0.988. Pairwise AUC comparisons indicated significant advantages of LGB-C over BC, GBDT, LR-C, RF-C, SC, and XGB-C, with AUCs ranging from +0.003 to +0.036, all of which were statistically significant (q < 0.05). These results highlighted LGB-C as the optimal model for LastPaAST prediction, with both strong overall discrimination and calibrated threshold performance. VC and CAT-C also showed competitive results and could serve as alternatives for deployment.

#### 3.3.2 Regression tasks

In CRR prediction (Fig. 4a), CAT-R and VR were closest to the observed values on the Taylor diagram: CAT-R achieved a centered root mean square difference (cRMSD) of 0.166, a correlation coefficient (r) of 0.885 (R^2^ ≈ 0.78), and a σ-ratio of approximately 0.880. VR followed with cRMSD = 0.168, r = 0.884 (R^2^ ≈ 0.78), and σ-ratio ≈ 0.850. XGB-R ranked third with cRMSD = 0.175, r = 0.873, and σ-ratio ≈ 0.900. The other models exhibited progressively higher cRMSD values. The Nemenyi post-hoc test showed significant differences between CAT-R and SR (*p* = 0.02) and between VR and SR (*p* = 0.01), confirming CAT-R and VR as the top-performing models for ARR prediction (Fig. 5a).

**Fig. 4.**
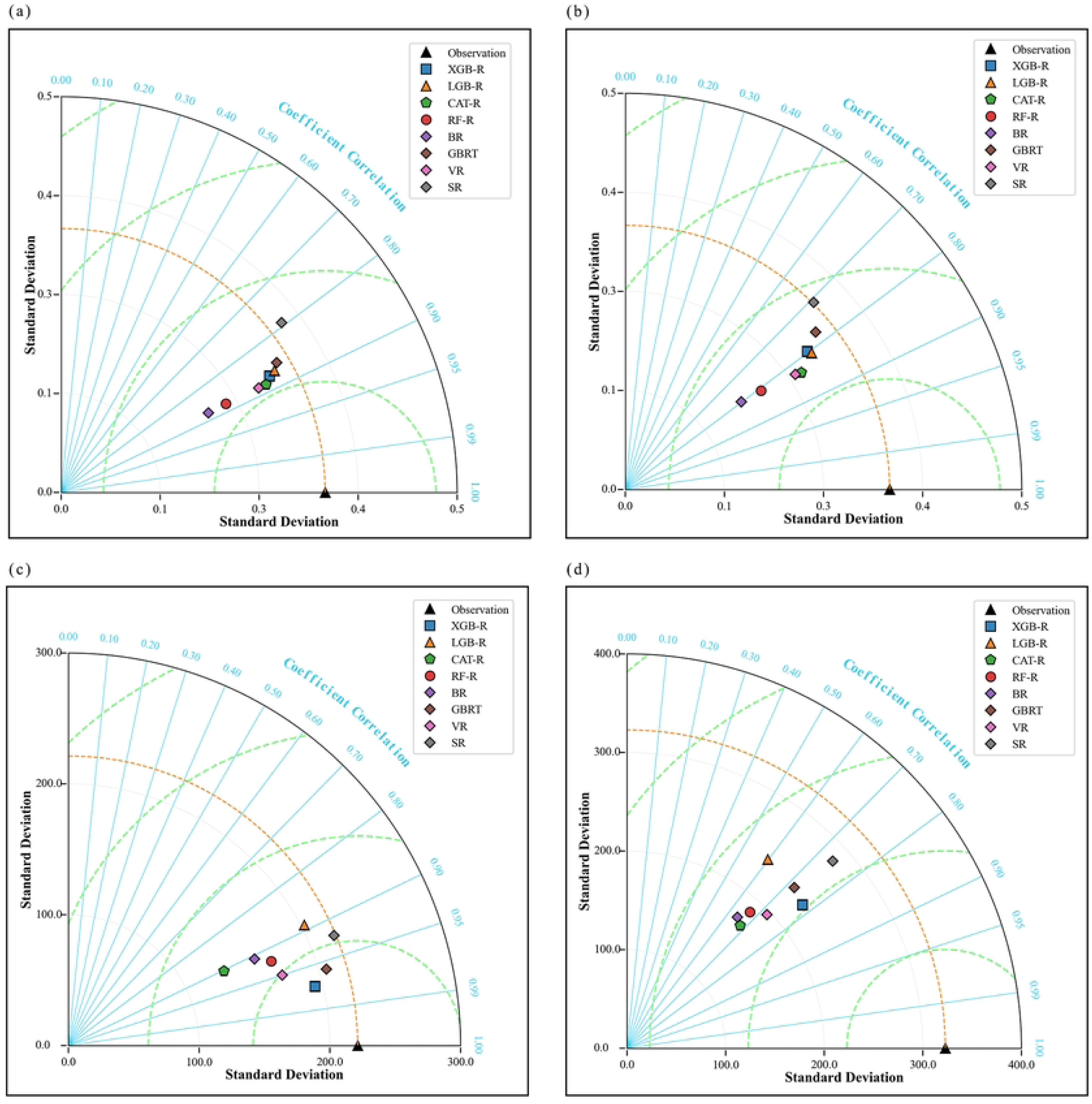
Taylor diagrams summarizing regression performance across four outcomes. (a) CRR; (b) CRR-PA–Last2; (c) ICU-LOS; (d) ICU-Death interval. The radial distance represents the modeled standard deviation, the azimuthal angle shows the Pearson correlation with observations, and the gray semicircles depict the centered root mean square difference (cRMSD). The orange dashed arc represents the standard deviation of the observed values.

**Fig. 5.**
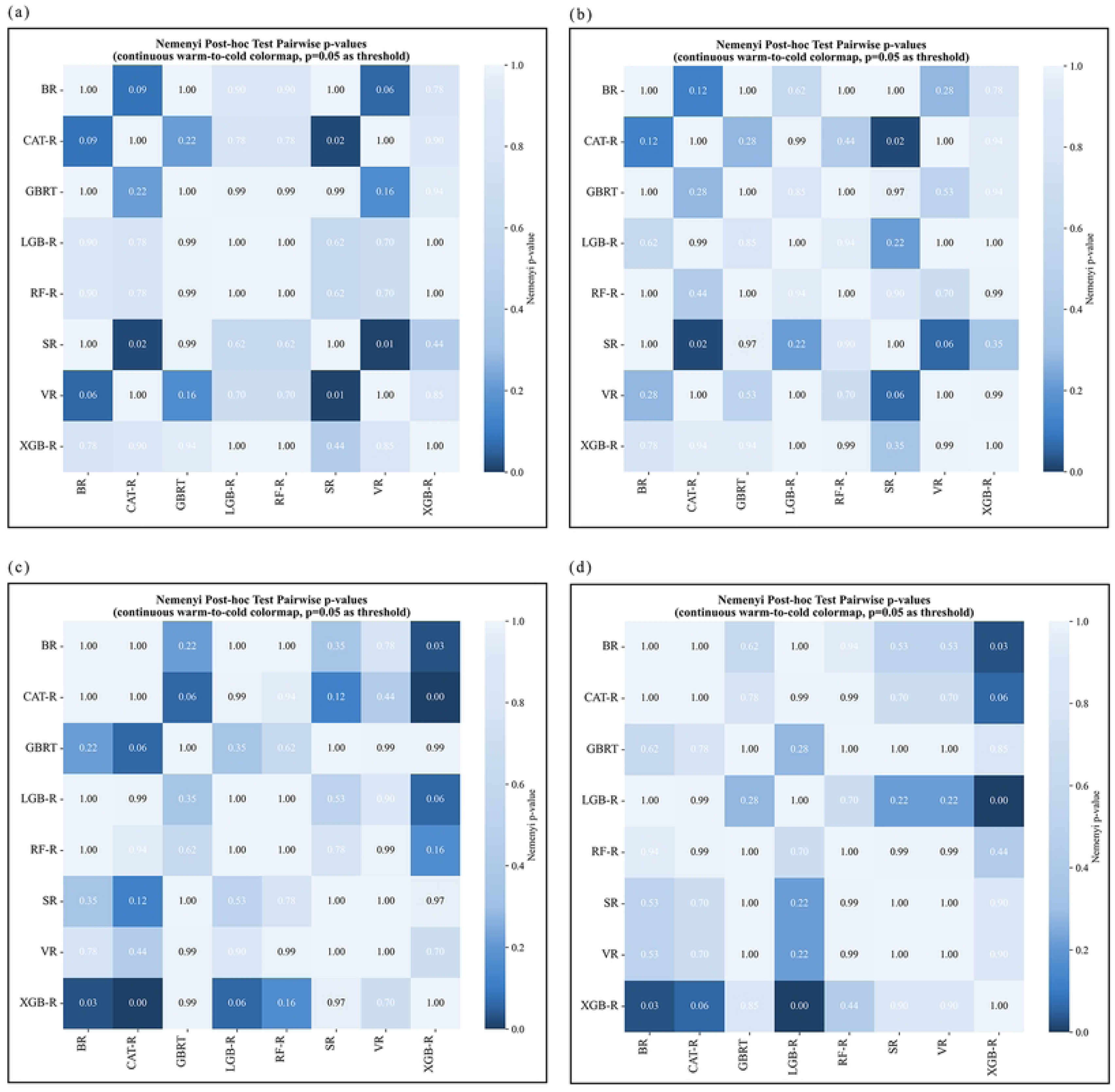
Pairwise Nemenyi post-hoc p-values for eight regressors. Heat maps display false-discovery rate-adjusted p-values comparing model ranks for (a) CRR, (b) CRR-PA–Last2, (c) ICU-LOS, and (d) ICU-Death interval. Darker blue tiles indicate smaller p-values, with cells showing *p* < 0.05 highlighted in deep blue and annotated in white.

In the CRR-PA-Last2 evaluation (Fig. 4b), the predicted standard deviations of all models converge on the observed value, forming a compact, nearly superimposed cluster of points on the Taylor diagram. The Friedman test revealed no overall differences (p > 0.05). The Nemenyi test revealed a significant difference only between BR and XGB-R (*p* = 0.03); all other comparisons were non-significant (*p* > 0.05), suggesting that XGB-R offered better-than-average computational efficiency while maintaining predictive accuracy, and the other models performed similarly (Fig. 5b).

For ICU-LOS prediction (Fig. 4c), XGB-R achieved the best overall performance with r = 0.972, cRMSD = 55.70 days, and σ-ratio ≈ 0.877 (194.03/221.27). GBRT followed with r = 0.959, cRMSD = 62.93 days, and σ-ratio ≈ 0.930. VR and RF-R showed systematic underestimation (σ-ratio ≈ 0.778 and ≈ 0.759), and CAT-R notably underestimated variability (σ-ratio ≈ 0.596). The Nemenyi test indicated that XGB-R significantly outperformed BR (*p* = 0.03) and CAT-R (*p* < 0.01), with no significant differences between the remaining models (*p* > 0.05), highlighting the balance between accuracy and amplitude alignment (Fig. 5c).

In predicting ICU-Death interval (Fig. 4d), SR performed robustly with cRMSD ≈ 221.16 days, r = 0.740, and σ-ratio ≈ 0.874 (189.28/322.87). However, XGB-R maintained the best overall balance with a lower cRMSD = 205.07 days, σ-ratio ≈ 0.712, and r = 0.774. The Nemenyi test showed significant advantages of XGB-R over BR (*p* = 0.03) and CAT-R (*p* < 0.01) with no statistically significant differences between the other models (*p* > 0.05), further confirming the superiority of XGB-R for tasks with heavy-tailed distributions (Fig. 5d).

Table 4 summarizes the mean ± SD and average ranks for each model across the four outcomes. XGB-R ranked first overall (Avg Rank = 1.95), demonstrating the lowest overall error (MSE 9.76 ×10^3^ ± 18.23, RMSE 64.11 ± 84.06, MAE 25.24 ± 32.20) and highest average R^2^ (0.77 ± 0.16).

**Table 4.**
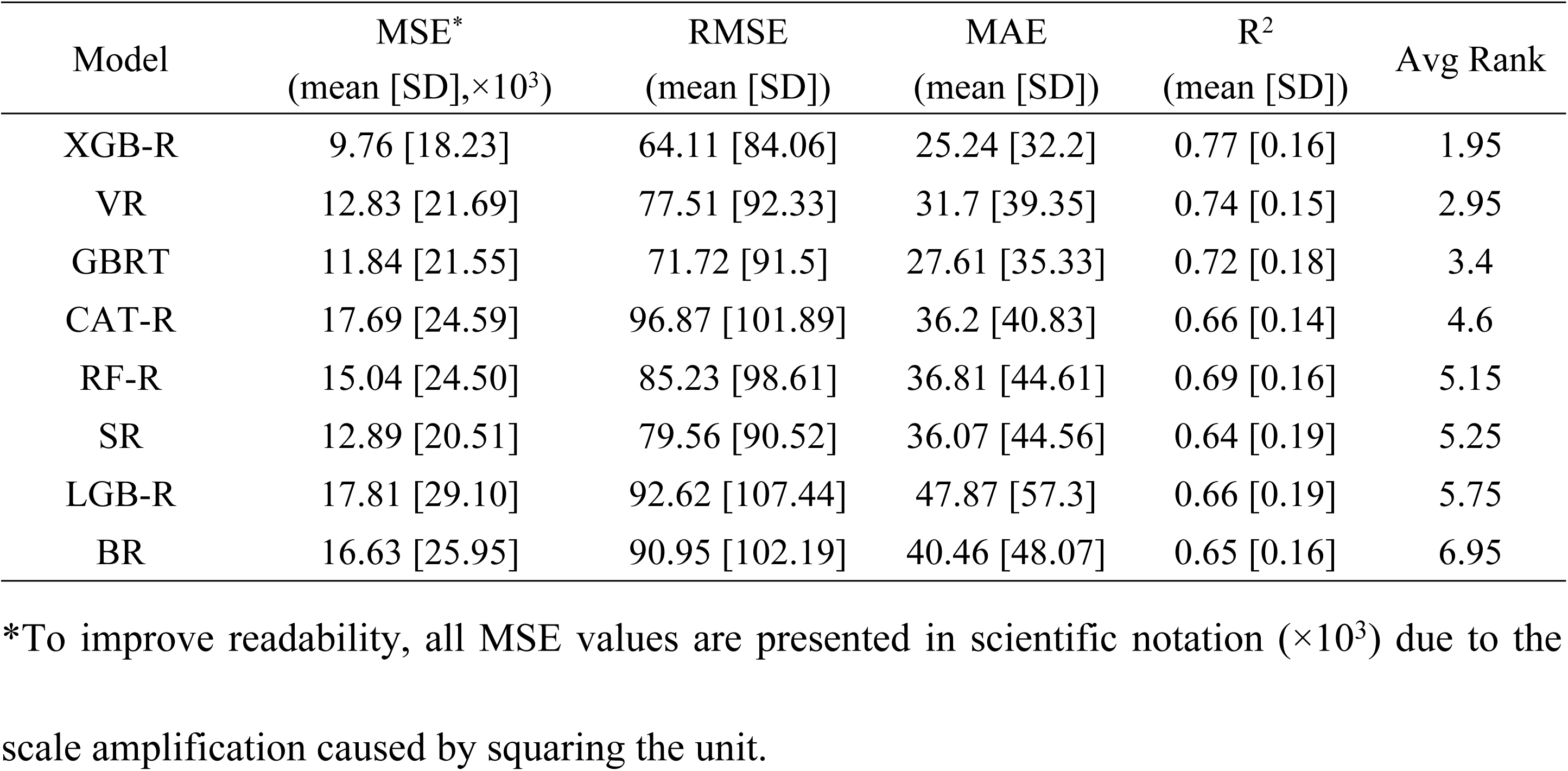
Multi-outcome regression performance and Friedman average ranks of the eight candidate models (mean ± SD)

VR (2.95) and GBRT (3.40) followed in second and third places, although VR and GBRT systematically under and overestimated the amplitude, respectively. CAT-R (4.60) was ranked fourth for its stability in ARR and Combined-ARR predictions. RF-R (5.15) and SR (5.25) ranked fifth and sixth, respectively, whereas LGB-R (5.75) and BR (6.95) ranked the lowest. Overall, XGB-R, VR, and GBRT demonstrated the best balance between the performance and computational efficiency across multiple outcomes.

### 3.4 Feature interpretation

#### 3.4.1 Classification tasks

##### (1) IHM

As shown in Fig. 6, the SHAP analysis of the LGB-C and RF-C models revealed that patient age (Age [years]), TAMT, PMX (ATC/DDD per DOT), UC-days, CARB (ATC/DDD per DOT), CARB (ATC-DDD), and CVC-days were the top contributors to the prediction of IHM. Age was the highest factor in both models, indicating a strong association with poor outcomes in older adult patients. Indicators of antimicrobial exposure and invasive procedures (e.g., combination therapies, urinary catheterization, and central venous catheterization) significantly influenced the model predictions. In particular, the high SHAP values for polymyxin and carbapenem use (regardless of whether measured by ATC/DDD per DOT or total ATC-DDD) highlight their predictive value for mortality risk. These features may partially reflect the severity of the underlying disease and treatment intensity rather than direct causality.

**Fig. 6.**
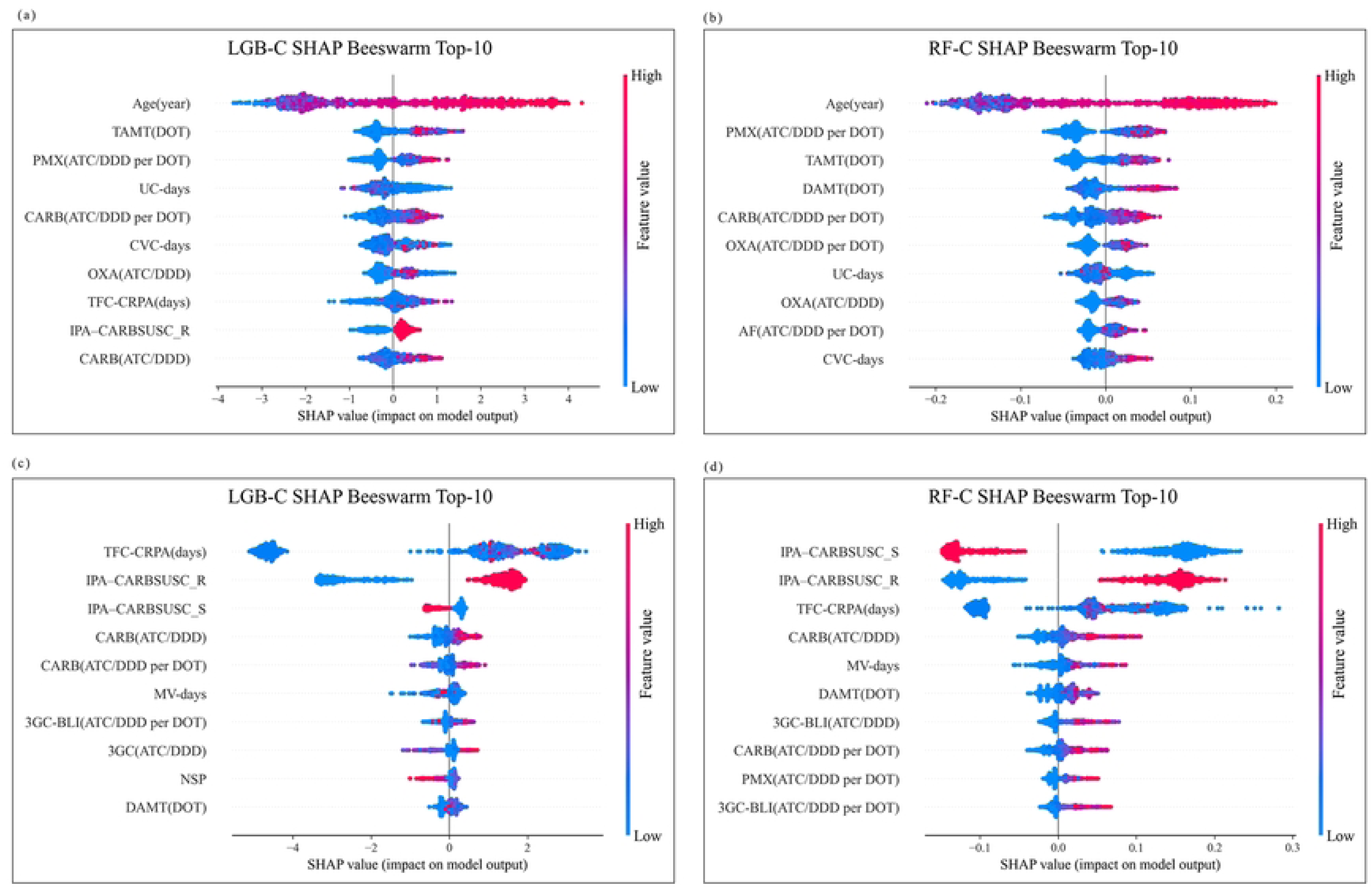
Global SHAP beeswarm plots for two binary outcomes. (a) LGB-C model for IHM; (b) RF-C model for IHM; (c) LGB-C model for LastPaAST; (d) RF-C model for LastPaAST. Each dot represents an individual SHAP value, with the color indicating a normalized feature value (blue = low; red = high). The features were ranked by the mean absolute SHAP value (|SHAP|), reflecting their relative contributions to the prediction. Only the top 10 features are shown; the SHAP values for all the remaining features are close to zero.

**Fig. 7.**
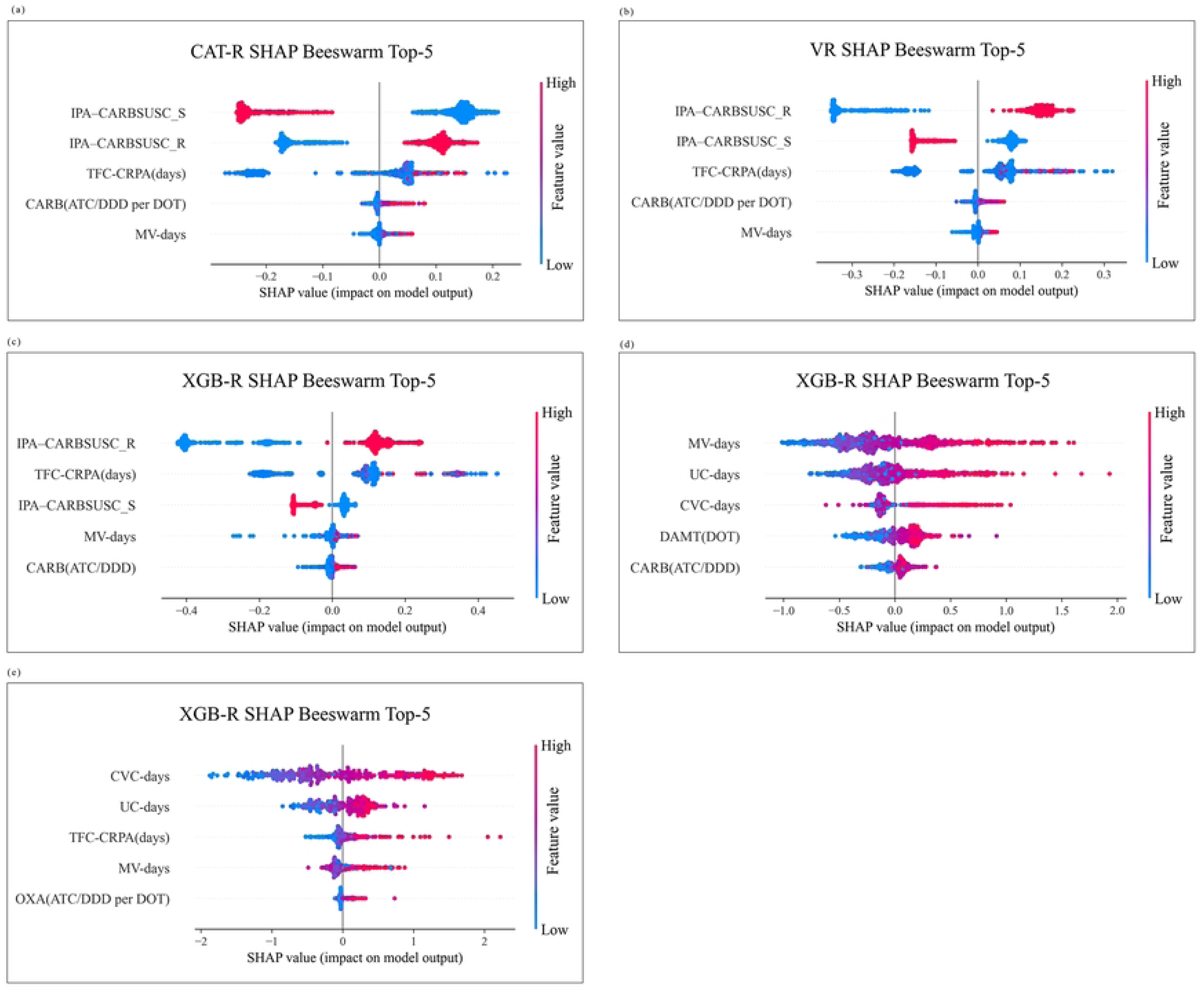
Global SHAP beeswarm plots for four regression outcomes. (a) CAT-R model for CRR; (b) VR model for CRR; (c) XGB-R model for CRR-PA–Last2; (d) XGB-R model for ICU-LOS; (e) XGB-R model for ICU-Death interval. Each plot shows the top 10 most influential predictors ranked by the mean absolute SHAP value (|SHAP|). Each dot represents an individual SHAP value, with the color encoding the normalized feature value (blue = low, red = high). Positive SHAP values indicate a higher predicted risk, whereas negative values indicate protective effects. The SHAP values for all remaining features are close to zero.

##### (2) LastPaAST

For the LastPaAST outcome, the SHAP results showed that variables closely related to TFC-CRPA and IPA-CARBSUSC dominated the feature rankings in both the LGB-C and RF-C models. In addition, exposure to carbapenems and third-generation cephalosporin/β-lactamase inhibitors (CARB [ATC/DDD and ATC/DDD per DOT], and 3GC-BLI [ATC/DDD and ATC/DDD per DOT]) exhibited high explanatory power. Other highly ranked factors included MV-days, NSP, and combination therapy duration, suggesting that invasive procedures and antimicrobial exposure are the key drivers of the development of carbapenem resistance in *P. aeruginosa*.

Across both the Death and LastPaAST outcomes, the feature importance rankings for LGB-C and RF-C were highly consistent. Age, antimicrobial exposure (especially to carbapenems and polymyxins), combination therapy, and invasive procedures have been consistently emphasized.

#### 3.4.2 Regression tasks

##### (1) CRR

In SHAP value rankings for CRR prediction, both CAT-R and VR models identified IPA-CARBSUSC_S and IPA-CARBSUSC_R as the two most important features, with average absolute SHAP values of 0.24 and 0.21 for CAT-R, and 0.28 and 0.25 for VR, respectively. These values are substantially higher than those of all other variables. TFC-CRPA (days) ranked third in CAT-R (SHAP = 0.18) and highest in VR (SHAP = 0.16). CARB (ATC/DDD per DOT) ranked third and fourth for CAT-R and VR, respectively, with SHAP values of 0.15 and 0.14. MV-days ranked fourth in CAT-R (SHAP ≈ 0.13) and fifth in VR (SHAP ≈ 0.12). All other candidate features had SHAP values below 0.10, with consistent rankings across both models. Overall, the SHAP analysis demonstrated high consistency between models, with IPA–CARBSUSC_S/R, TFC-CRPA, CARB exposure, and MV-days identified as the top four contributors to CRR prediction.

##### (2) CRR-PA–Last2

In the CRR-PA–Last2 prediction, SHAP rankings from the CAT-R and XGB-R models similarly emphasized the dominant role of resistance status and combination therapy duration. For CAT-R, IPA–CARBSUSC_S had the highest mean SHAP value (0.1716), followed by IPA– CARBSUSC_R (0.1232), TFC-CRPA (0.0916), CARB (ATC/DDD per DOT, 0.0079), and MV-days (0.0066). For XGB-R, IPA–CARBSUSC_R ranked first (0.2053), followed by TFC-CRPA (0.1461), IPA–CARBSUSC_S (0.0544), MV-days (0.0145), and CARB (ATC/DDD, 0.0084).

Both models confirmed that early resistance and prolonged multidrug exposure were decisive predictors, whereas MV-days and carbapenem dose made relatively modest contributions.

##### (3) ICU-LOS

SHAP analysis from the XGB-R model indicated that ICU length of stay (log-transformed) was primarily driven by five features. MV-days had the highest average SHAP value (0.384) with a wide and positively skewed distribution, indicating a strong marginal impact. UC-days ranked second (SHAP = 0.262), followed by CVC-days (SHAP = 0.183), both of which had a positively skewed SHAP distribution. The DAMT DOT ranked fourth (SHAP = 0.157), with contributions on both sides of the axis but a predominance of positive values. CARB (ATC/DDD) ranked fifth (SHAP = 0.073) with a modest but consistent positive influence. All other features had SHAP values < 0.06. These contributions were visualized using beeswarm plots, which clearly illustrated the marginal effects of each feature on the log-transformed ICU-LOS prediction. A note in the figure caption indicates that log transformation was applied throughout the modeling and interpretation processes owing to the strong skewness in the outcome distribution.

##### (4) ICU-Death intervals

In the SHAP analysis of the XGB-R model for the log-transformed ICU-Death interval, CVC-days had the highest average SHAP value (0.703) with a wide and positively skewed distribution, indicating its dominant marginal effect. UC-days followed (SHAP = 0.268), indicating a potential association with increased predicted mortality risk, which may reflect the impact of prolonged urinary catheterization. TFC-CRPA ranked third (SHAP = 0.167), suggesting a positive correlation between prolonged exposure to MDR pathogens and longer time-to-death. MV-days and OXA (ATC/DDD per DOT) ranked fourth (SHAP = 0.150) and fifth (SHAP = 0.054), respectively. While MV-days showed mostly positive contributions, the influence of OXA was modest but concentrated in high-value regions. All other features had SHAP values of < 0.047. These results were visualized in beeswarm plots showing the marginal contributions of each feature, and the captions indicate that log transformation was applied consistently across model training, prediction, and interpretation.

Due to the strong skewness in the ICU-LOS and ICU-Death interval distributions, both outcomes were log-transformed, consistently modeled, and interpreted on a log scale. Other continuous variables were imputed using medians and standardized via Z-score transformation. Categorical variables were imputed using modes and one-hot encoding. All transformations were implemented within a unified pipeline and transformed target regressor structure to ensure reproducibility. The final beeswarm plots display the five features with the highest marginal contributions to the prediction of (log-transformed) ICU outcomes.

## 4 Discussion

We innovatively developed and validated a hybrid AutoML ensemble framework based on a retrospective single-center ICU cohort. The pipeline integrates conventional learners such as logistic regression and Random Forest with gradient-boosting algorithms including XGBoost, LightGBM, and CatBoost. Within a unified preprocessing, training, cross-validation, and ensemble pipeline, the framework simultaneously achieved quantitative regression predictions of continuous outcomes, such as CRR, ICU length of stay, and time-to-death intervals, as well as binary predictions for categorical outcomes, such as in-hospital all-cause mortality and antimicrobial susceptibility of the last isolated strains. Compared to previous research focusing solely on individual tasks or requiring separate maintenance of multiple models, this integrated approach significantly reduced clinical deployment and maintenance complexity and surpassed prior ICU AMR prediction outcomes across several metrics [34–36]

In binary classification tasks, fixing the sensitivity at 0.80 facilitated a more intuitive comparison of the trade-off between false-positive rates and the overall discriminative capacity. The VC achieved the highest AUC for mortality risk prediction, making it particularly suitable for scenarios requiring maximal detection coverage. Similar to the findings of Chhillar and Singh, who reported an AUC of 0.9985, accuracy of 99.42%, and sensitivity of 99.20% by integrating SVM, XGBoost, and MLP models on the WDBC breast cancer dataset, our study confirmed the superior predictive capability of soft voting ensembles for mortality risk [37]. Additionally, the Random Forest Classifier (RF-C) demonstrated optimal performance in predicting sepsis severity, mortality, and ICU length of stay (ICU-LOS) [38]. Our analysis further validated RF-C as the preferred model when minimal false-positive rates were clinically essential. To predict the susceptibility of the last isolated strain, LGB-C combined the highest AUC with the optimal threshold performance, making it the recommended first choice. Moreover, VC and CAT-C exhibited strong performance in multi-model ensembles, providing diversity and weighted optimization.

In the regression tasks, the XGB Regressor (XGB-R) consistently demonstrated the lowest RMSE and best average rank performance, aligning with the systematic review by Almeida et al. on hospital length of stay prediction, where XGBoost outperformed traditional methods, particularly in resource-sensitive contexts requiring minimal overall prediction errors [39]. The SR and gradient-boosted regression trees with Huber loss (GBRT-Huber) showed robust performance at extreme quantiles, consistent with Wang’s robust boosting theory, underscoring the suitability for predicting heavy-tailed distributions such as time-to-death [40]. CatBoost Regressor (Cat-R), with balanced accuracy and interpretability, is ideal to handle mixed-feature scenarios. Collectively, gradient-boosting methods provided the best compromise across correlation, amplitude matching, and error scale, whereas the traditional Bagging and Random Forest approaches were less suitable owing to systematic amplitude biases. The medical imaging findings of Srinivasu et al. support this conclusion, highlighting CatBoost combined with SHAP as highly discriminative yet transparent [41].

SHAP analysis was integrated into our model interpretability workflow to achieve a comprehensive explanatory loop in line with the Explainable AI (XAI) framework proposed by Bilal et al. [42]. By visualizing the global feature importance alongside individual decision pathways, the analysis clarified the clinical logic inherent to model predictions and provided actionable quantitative evidence supporting antimicrobial stewardship and life-support adjustments.

Global SHAP explanations validated the model’s sensitivity to well-established clinical risk factors: increased age, prolonged mechanical ventilation, and central venous catheterization were positively correlated with mortality risk (Fig. 6). This finding aligns closely with the established practice of incorporating age and intensity of organ support into the traditional APACHE II scoring system [43]. IPA-CARBSUSC_R demonstrated significant predictive contributions to mortality risk in both LGB-C and RF-C, with mean absolute SHAP values of 0.45 and 0.42, respectively, highlighting the importance of timely AST and precise medication administration [44]. Additionally, CARB ATC/DDD and combined drug-use days ranked prominently in the global SHAP analyses of CAT-R and VR, reinforcing that widespread carbapenem use is related to increased resistance in *P. aeruginosa* [45–47]. XGB-R further identified interactions between “time from admission to event,” dynamic support indicators (MV-days, CVC-days), and cumulative medication intensity, providing quantitative evidence to inform personalized interventions.

Furthermore, combination antibiotic regimens involving DAMT and PMX administration positively influenced mortality and last antibiotic-susceptibility outcomes, reflecting the intensity of critical infection therapy but also possibly indicating negative impacts, such as antibiotic-associated dysbiosis or toxicity. PMX exposure influenced both IHM and susceptibility outcomes.

Dosage analysis revealed that polymyxin B accounted for a cumulative consumption of 1,270.78 ATC/DDD (217.09 ATC/DDD per DOT), representing 55% of total polymyxin consumption at the study site—significantly higher than polymyxin E and colistin sulfate formulations. Ecological studies from Brazilian ICUs similarly reported significant correlations between rising annual polymyxin B consumption and the incidence of carbapenem/polymyxin co-resistant *P. aeruginosa* (β = 0.71, p < 0.01) [48]. Research on the molecular mechanisms further indicated that PMX-induced lipid A remodeling and mutations in the pmrAB and phoPQ two-component regulatory systems reduce outer membrane permeability, restrict carbapenem entry, and promote cross-resistance. These findings suggest that although polymyxin B remains a critical “last-line” treatment against MDR *P. aeruginosa*, the intense use thereof also exacerbates selection pressure for resistance. Meanwhile, the membrane-disruptive combination therapy of polymyxin B and meropenem has demonstrated potential *in vitro* and animal studies to inhibit heterogeneous resistance, providing a foundation for future precision dosing and combination therapy strategies [49]. In addition to pharmaceutical factors, advanced age, extended duration of mechanical ventilation, and central venous catheterization correlated positively with mortality risk, indicating declining physiological reserves and heightened complication risks, consistent with the established APACHE II and SOFA prognostic scoring systems.

To assess real-time applicability further, models were trained and evaluated on a standardized hardware platform (Intel® 13th-Gen Core™ i5-13400F processor with 10 cores/16 threads, base 2.5 GHz, turbo up to 4.6 GHz, and 32 GB DDR4 RAM) under a consistent software environment (Python 3.8; scikit-learn 1.1.2; XGBoost 1.5.0; LightGBM 3.3.2; CatBoost 1.0.5), with single-prediction latency measurements. The results demonstrated that classification tasks using a VC and SC averaged inference times below 50 ms (Table 5). Regression tasks employing the VR and SR averaged between 40 and 50 ms, comparable to single-model baselines such as GBRT, XGB, and LGB (Table 6). Despite longer inference times for the Bagging models, voting and stacking ensembles maintained millisecond-level responsiveness, thus satisfying real-time ICU clinical decision-support requirements.

**Table 5.** Average prediction time of classification models.

| Model | Average Prediction Time (ms) |
| --- | --- |
| LR-C | 3.80 |
| GBDT | 6.50 |
| XGB-C | 8.80 |
| LGB-C | 12.30 |
| CAT-C | 14.10 |
| RF-C | 61.80 |
| VC | 46.40 |
| SC | 45.50 |
| BC | 719.30 |

**Table 6.** Average prediction time of regression models.

| Model | Average Prediction Time (ms) |
| --- | --- |
| GBRT | 4.30 |
| XGB-R | 7.60 |
| CAT-R | 8.00 |
| LGB-R | 8.90 |
| RF-R | 61.70 |
| VR | 40.80 |
| SR | 37.00 |
| BR | 596.90 |

Tables 5 and 6 summarize the average single-prediction inference times of the classification and regression models, respectively.

Despite the significant findings, this study has limitations. First, its retrospective single-center design necessitates multicenter prospective validation for better generalization. Second, the threshold choices in VTF-MI-L1 methods involve subjective empirical decisions, potentially requiring recalibration for specific scenarios. Finally, the computational complexities of AutoML and cross-validation pose challenges to resource management and real-time deployment, suggesting that future efforts should balance model complexity with practical responsiveness.

## 5 Conclusion

This retrospective single-center cohort study presented a novel hybrid AutoML ensemble framework combining traditional learners (e.g., logistic regression, Random Forest) with gradient-boosting algorithms (XGBoost, LightGBM, and CatBoost). The framework achieved precise prediction and highly interpretable outputs for various ICU outcomes, including CRR, susceptibility to the last isolated strains, IHM, ICU stay duration, and time-to-death. Future studies should emphasize multicenter prospective validation and model optimization to improve generalization and real-time clinical decision support.

## Data Availability

All relevant data are within the manuscript and its Supporting Information files.

## Supporting information

**S1 File. Patient-Level Filtered Clinical and Antimicrobial Exposure Features Dataset.** It includes detailed patient characteristics such as Patient ID, Age(year), sex, and various metrics related to antimicrobial use (e.g., DAMT(DOT), CAMT(DOT), TAMT(DOT), AG(ATC/DDD per DOT), FQ(ATC/DDD)), as well as clinical exposure indicators (e.g., CVC-days, MV-days, UC-days). The dataset also includes information on antimicrobial susceptibility (e.g., IPA– CARBSUSC) and other relevant clinical parameters.

S2 File. This is the Clinical Outcome Indicators Dataset. This file contains the primary and secondary clinical outcome variables for each patient, aligned with the Patient ID in the ‘Patient-Level Filtered Clinical and Antimicrobial Exposure Features Dataset’ (S1 File). Key outcome indicators include IHM, LastPaAST, CRR, ICU-LOS(days), and ICU-Death Interval (days).

